# Compliance with infection prevention and control practices and associated factors among healthcare workers in Tanzania: Experience from Katavi Regional Referral Hospital in Katavi

**DOI:** 10.1101/2025.11.06.25339732

**Authors:** Cesilia Charles, Lutengano Mkonongo, David Masanja, Pendo Edward, Damian Maruba, Philipo Felix Mwita, Baraka Nkondo, Frank Elisha, Edward Bucheye, Abel Nyika, Avent Kalumiana, Emmanuel Amsi, Elly Ambikile, Nathanael Sirili, Joshua Mollel, Bernard Njau, Radenta Bahegwa, Deogratias Banuba

## Abstract

**Introduction:** Compliance with Infection Prevention and Control practices remains a key challenge, affecting the safety of both patients and healthcare workers. Poor compliance raises the risk of Hospital-Acquired Infections (HAIs), antimicrobial-resistant organisms, and hinders efforts to provide safe healthcare by 2030.

**Objective:** This study aimed to evaluate compliance levels and factors associated with infection prevention and control practices among HCWs at Katavi Referral Regional Hospital in Tanzania.

**Methods:** A hospital-based cross-sectional study involving 195 healthcare workers was conducted between 24^th^ July and 23^rd^ August 2025. Questionnaires and observation checklists were used to collect sociodemographic data, individual-level, hospital-level factors, and the availability of IPC supplies. A validated Compliance with Standard Precautions Scale (CSPS) tool developed by WHO was used to measure compliance levels. Data were analysed with the aid of STATA version 15.0, using bivariate and multivariate modified Poisson regression models to assess factors associated with IPC compliance.

**Results:** The study revealed that the overall compliance with IPC practices was 68.9% with 39% of HCWs having a high overall compliance with IPC practices, scoring <80%, and 61% had low compliance, scoring >80%. Also, factors significantly associated with compliance with IPC practices were doctor profession (APR: 0.32;95% CI: 0.19, 0.57), blood/body fluid exposure (APR: 1.55;95% CI: 1.095,2.19), motivation at workplace (APR: 1.43;95% CI: 1.02,2.02), supportive supervision (APR:1.92;95% CI: 1.09,3.38), and presence of IPC committee (APR:1.61;95% CI:1.07,2.40). The most common available IPC supplies were HH items, PPEs, and waste management items (100%). However, some IPC supplies were unavailable, including water and soap in latrines.

**Conclusion:** Overall compliance with IPC practices among HCWs was suboptimal, with only 39% achieving the recommended level of >80%. Compliance was positively influenced by exposure to body fluids, workplace motivation, supportive supervision, and the presence of an IPC committee. While essential supplies such as hand hygiene items, PPEs, and waste management items were available, gaps remained, particularly the lack of water and soap in latrines. To increase compliance levels, the health system should strengthen IPC activities within the IPC committee and address resource gaps within the health facilities.

## Introduction

Global initiatives by the United Nations (U.N.) to promote healthy lives and well-being for all populations emphasize a range of health targets, including universal health coverage (UHC), which focuses on the right to health, financial risk protection, and access to quality health services [1]. According to the World Health Organization (WHO), quality healthcare is regarded as patient-centred, safe, effective, timely, efficient, and equitable [3]. Within this framework, infection prevention and control (IPC) is recognized as a fundamental element in delivering high-quality care by addressing HAIs, antimicrobial resistance (AMR), and emerging pathogen containment [5–7]. IPC practices are designed to protect patients, staff, and visitors from exposure to disease-causing agents (germs) that may be present in the healthcare environment or HAIs [4]. It includes hand hygiene, PPE use, handling of sharp devices, immunization, post-exposure prophylaxis, and isolation enacted for the prevention of HAIs [8].

Hospital-acquired infection (HAI) is the most common adverse event in healthcare delivery systems worldwide, threatening the health of patients and healthcare workers [11–13]. A global report on infection prevention and control in healthcare settings shows that HAIs kill 7 patients in high-income countries and 15 patients in LMICs annually [8]. In Sub-Saharan African countries, the HAI rate is 12.9% of which the highest burden (19.7%) was observed in East Africa [17–21]. Likely in Tanzania, the rate of HAI is 14.8% higher than in the developed world [14]. The main cause for the high HAI rate is non-compliance with IPC practices among HCWs. Additionally, the lack of an IPC committee, IPC training, and IPC policies/guidelines impedes their compliance [22–24].

Fortunately, 55–70% of HAIs can be prevented using available evidence-based IPC strategies. [16]. Regrettably, only 34% of WHO member states in 2021 had a fully implemented IPC programme across their country, and among them, only 19% reported having a system in place to monitor the effectiveness and compliance with the implemented prevention and control activities [17]. Bridging the IPC compliance poses a significant risk for mental health disorders, such as anxiety, depression, adjustment disorder, panic attacks, post-traumatic stress disorder, and economic burden to the health systems and family as a result of HAI and AMR[15].

In Tanzania, the IPC of healthcare workers in healthcare facilities has been addressed since 2004, and a new version of the national IPC documents was published in July 2018, corresponding with WHO recommendations [25]. These guidelines have been reformulated to improve the compliance of HCWs with IPC precautions and to interrupt the circulation of infection, consequently protecting the HCWs, patients, and the community from HAIs and their consequences. Despite the interventions, the implementation of the IPC recommendation is still far from satisfactory in most hospital settings, with a direct impact on the safety of patients and healthcare workers. Therefore, this study aims to assess the compliance with IPC guidelines and associated factors among HCWs to generate evidence-based strategies that address the poor compliance and strengthen the resilient health system at healthcare facilities by enhancing preparedness and response to future unexpected infectious epidemics.

## Materials and Methods

### Study design

A facility-based cross-sectional study was conducted at Katavi RRH using a quantitative approach to assess the compliance levels with Infection Prevention and Control (IPC) and associated factors among HCWs.

### Study setting and period

Data were collected from 24^th^ July to 23^rd^ August 2025 at Katavi Regional Referral Hospital (KRRH), one of 28 Tanzanian government regional referral hospitals located in Mpanda Municipality. According to the 2022 Census, the hospital serves a catchment population of 1,152,958 (26). The hospital offers a range of inpatient and outpatient services, including medical, surgical, obstetrics and gynaecology, child health, investigation, and medication services. It receives patients from Katavi’s councils, neighbouring regions, and the Democratic Republic of Congo. It refers patients to the zonal hospitals in Mbeya (550 km), Bugando Hospital in Mwanza 745 km, KCMC Hospital in Kilimanjaro (1265 km), Muhimbili National Hospital (1500 km), and Benjamin Mkapa Hospital (712 km). KRRH has 445 employees of various cadres, which is below the minimum range of 481, a recommended staffing requirement for a Regional Referral Hospital according to Tanzania’s MoH guidelines [27]. Out of 445 employees, 366 (82.2%) are HCWs (i.e., 83 doctors, 252 nurses, and 31 laboratory practitioners).

### Study Population

All healthcare workers who were directly involved in patient care, including nurses, doctors, and laboratory practitioners.

### Exclusion and inclusion criteria

#### Inclusion criteria

- HCWs who are permanently employed at Katavi RRH with a minimum of six months’ experience in caring for patients during ward rounds, doing procedures, and having actual contact with the patients and their surroundings.
- HCWs who had the will to provide informed consent and participate in the study.

#### Exclusion criteria

- The study excluded volunteers, medical students, and participants who were on leave or unavailable during the data collection period.

#### Sample size determination

The sample size was estimated using the Kish-Leslie formula for sample size determination.

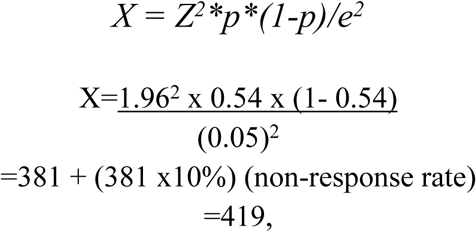

Where Z: Standard normal deviate (1.96), corresponding to a 95% confidence level, and e: a margin of error of 5% with the assumption of a 54% prevalence (p) of compliance with IPC practices [28]. The sample size was estimated to be 381. The researchers anticipated a 10% non-response rate. The final sample size was therefore estimated as 419.

Since the HCWs population (N=366) was known from the human resource registry and was smaller than the minimum sample size calculated from Kish’s formula, thus, the Cochran formula was used to adjust the sample size for a finite population of 366 HCWs.

n = N*X / (X + N – 1), where:

N is the population size (366).

X is the minimum sample size calculated by the proportional formula

Thus:

n = 366*419/ (419+366-1) = 195

The adjusted sample size was 195 HCWs (**Table 1),** with a 100% response rate.

**Table 1:** Adjusted sample size for equal representation of participants.

| Professions | Population per cadre | Percentage (%) | Proportional sample size |
| --- | --- | --- | --- |
| Doctors | 83 | 22.7 | $83/366 \times 195 = 44$ |
| Nurses | 252 | 68.8 | $252/366 \times 195 = 134$ |
| Laboratory practitioners | 31 | 8.5 | $31/366 \times 195 = 16$ |
| **Total** | **366** | **100** | **195** |

### Sampling procedures

HCWs were stratified into three categories: doctors, nurses, and laboratory practitioners. A list of the total number of HCWs (doctors, nurses, and laboratory practitioners) was obtained from the hospital’s human resources department. Using the calculated proportionate sample size of the respective stratum, respondents were recruited into the study using a random number table and then asked for informed consent to participate.

### Study variables

**Independent variables were:** social demographic characteristics, working experiences, years of working experience, history of needle stick injury, hepatitis B vaccination, supportive supervision, IPC training, IPC motivation at the working workplace, availability of IPC supplies, and presence of an IPC committee.

**The dependent variable was compliance with standard IPC practices,** defined as the extent to which HCWs follow IPC guidelines, protocols, standards, and policies designed by WHO to ensure patient safety, infection control, and ethical practices within the healthcare setting.

### Measurement of the dependent variable

The dependent variable was the overall compliance with IPC practices, which was dichotomous, categorized as either high compliance or low compliance. A four-point Likert scale (Never, Often, Sometimes, and Always) was used to measure the 20 items for IPC practices, including hand washing, waste management, decontamination, and the use of PPE. Items 2, 4, 6, and 15 were negatively stated, while the rest of the items were positively stated. A score of 1 was given to an “always” response in positively worded statements and “never” option in negatively worded statements, while 0 was given for the rest of the responses, giving a total possible range score of 0 to 20. The internal reliability score was Cronbach’s alpha =0.8, indicating acceptable internal reliability. The compliance was quantified as the mean percentage value derived from the 20 items. The dichotomous dependent variable was categorized as high compliance (80%) with an average score above 16 and low compliance (<80%) with an average score below 16. This categorization was based on Tanzania’s national guidelines for the recognition of the implementation status of quality improvement initiatives in health facilities, including IPC improvement initiatives, where an HCW compliance rate of at least 80% was considered the desired level of compliance [29].

### Data Collection

A validated, structured, pre-coded, and self-administered questionnaire adapted from the WHO IPC assessment framework [30] was used to collect sociodemographic characteristics, compliance with IPC practices, and factors that could potentially be associated with compliance among HCWs at KRRH. Data were collected using the Kobo Toolbox. The validity and reliability of the developed questionnaires were tested. Before data collection, a pre-test was conducted on 10% of the total sample outside the targeted hospital. Data were collected in accordance with the necessary ethical guidelines and protocols, following receipt of ethical approval from the Institutional Review Board of the Medical Research and Ethics Committee in Mbeya, Tanzania. Additionally, an observational checklist was developed to gather information on the availability of IPC supplies among HCWs in nine departments at KRRH.

### Data analysis

After completing the data collection using a Kobo Toolbox, the data were uploaded to STATA version 15.0 for cleaning and final analysis. Categorical variables were summarized and presented as frequencies and percentages, while continuous variables were summarized using means and standard deviations (SD). The chi-square test was performed to determine the association between independent variables and the dependent variable at a significance level of P-value ≤ 0.05. Variables with significant association and those reported in previous literature as factors associated with compliance with IPC practices were considered in a bivariate modified Poisson regression model, as the outcome for the study was more than 15%. Factors with p-value <0.2 in the bivariate modified Poisson regression analysis were added to the multivariate modified Poisson regression model to identify factors significantly associated with high compliance with IPC practices at p-value <0.05.

### Ethical consideration

The ethical clearance was obtained from the Institutional Review Board (IRB) of the Medical Research and Ethics Committee (MMREC) in Mbeya, Tanzania (SZEC-2439/ R.A/25/11). Permission to conduct the study was also secured from the Research and Publication department of Katavi Regional Referral Hospital (KRRH). Study participants provided written informed consent to participate voluntarily. The authors declared that the information presented in this research article is original work that has not been presented elsewhere.

## Results

### Demographic Characteristics of Healthcare Workers

Of 195 HCWs who participated in the research study, most 135 (69.2%) were nurses, while the least 17(8.7%) were laboratory practitioners. The median age of respondents was 31.6 (±5.5) years, with a male preponderance of 113 (57.9%). The majority of HCWs, 93 (47.7%), had a diploma-level education, and 86(44.1%) had worked between 2 and 5 years in healthcare service delivery at KRRH. Table 2 shows the baseline characteristics of the study participants.

**Table 2:**
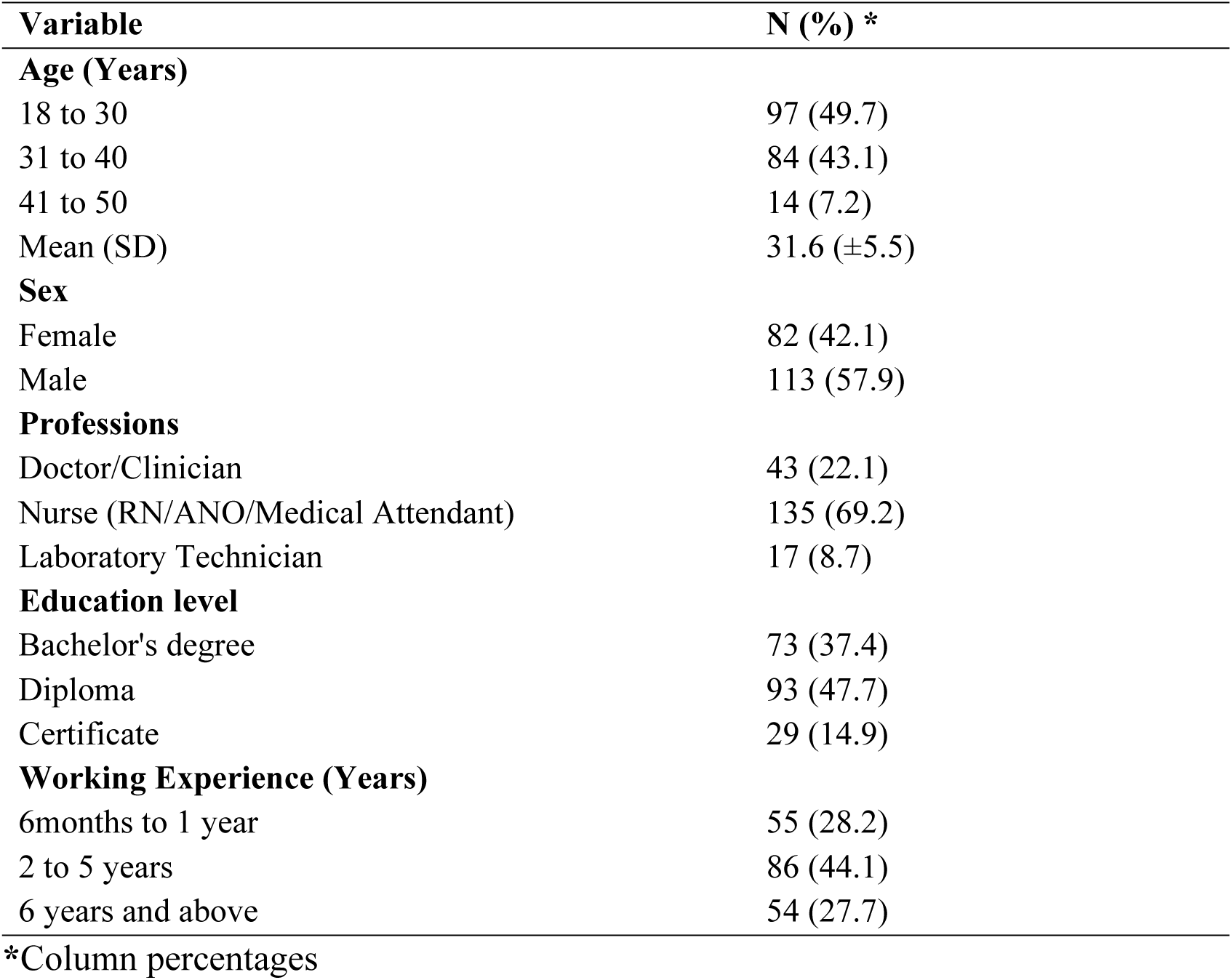
Baseline characteristics of the study participants (N=195)

### Compliance with the Infection Prevention and Control Standards precautions

The overall mean compliance with IPC among HCWs at KRRH was 68.9%. Out of 195 study participants, 39% (n=76/195) had a high overall compliance with IPC practices and scored an average of 16 (80%) or above. In contrast, 61% (n = 119/195) of HCWs had low compliance with IPC practices, scoring an average of less than 16 (80%). **Figure 1** presents the overall compliance status among HCWs at KRRH.

**Figure 1:**
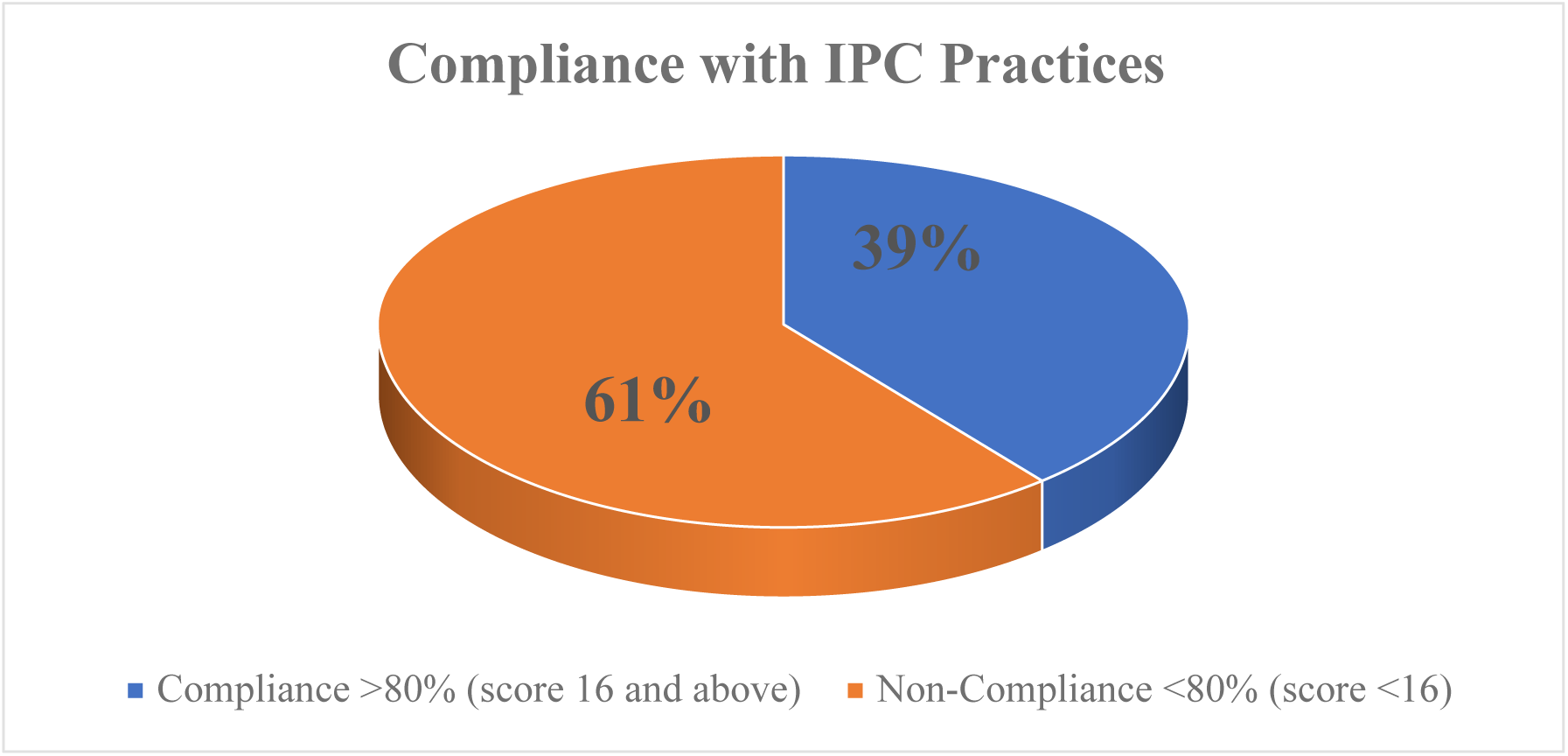
Overall Compliance status among HCWs at Katavi Regional Referral Hospital

### Compliance with IPC Standard Precautions

The majority of HCWs, 88.2% (172/195), reported the highest compliance with wearing gloves when exposed to body fluids, blood products, and patient excretion; however, more than half, 53.8%(n=105/195), of HCWs do not take a shower in case of extensive splashing, even after they have put on PPE. Nearly 68%(n=132/195) of HCWs use water alone for handwashing, and only 35.9% reported that it is incorrect for the sharps box to only be disposed of when it is full. **Table 3** represents the compliance with IPC precautions among HCWs at KRRH.

**Table 3:**
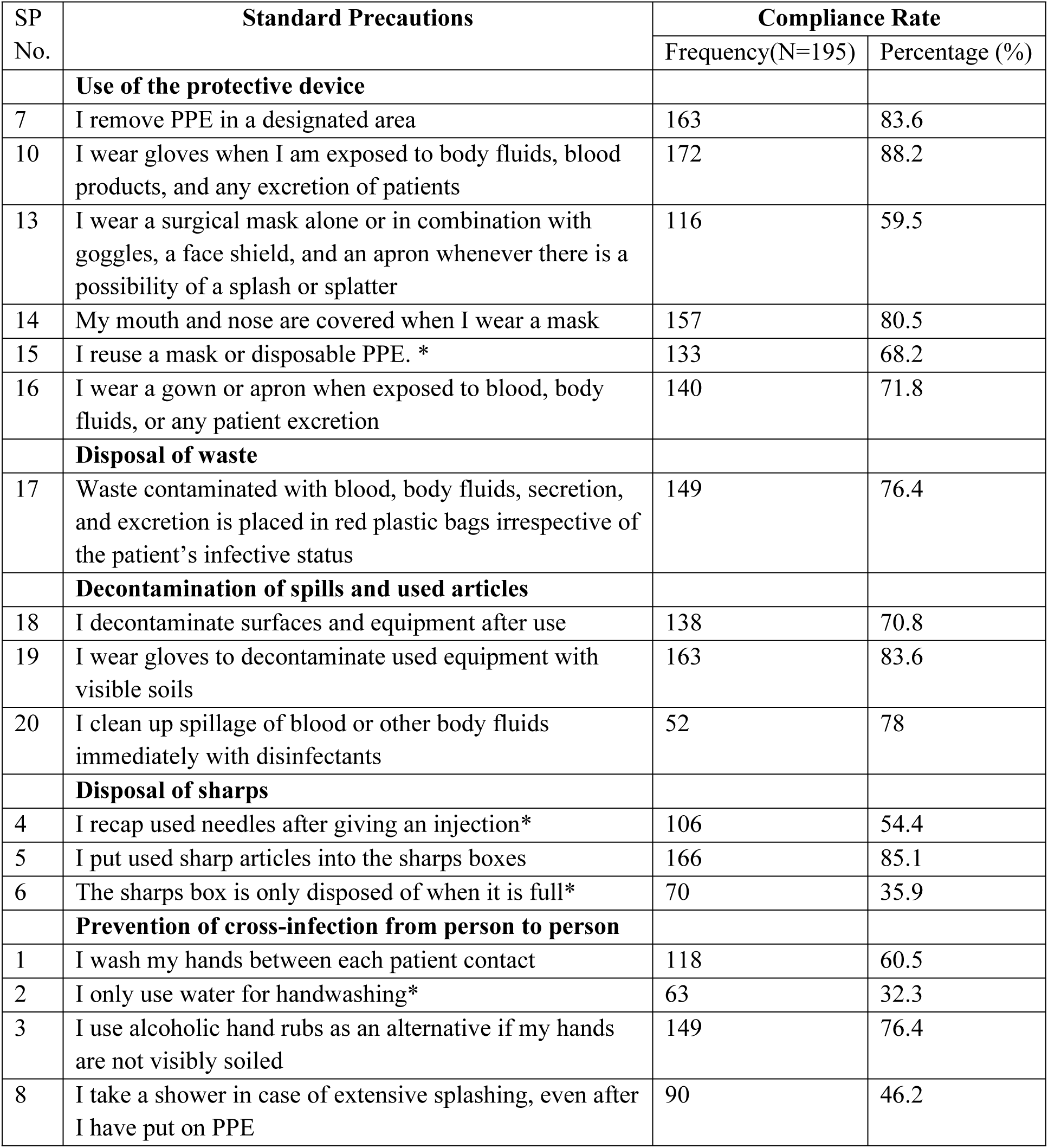
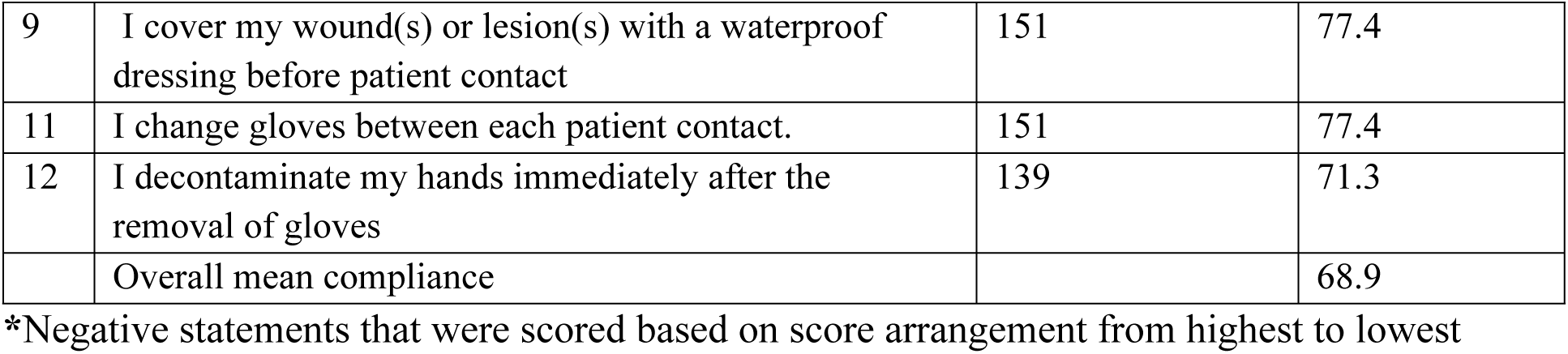
Compliance with IPC Precautions among healthcare workers (N=195), as measured by the WHO CSPS.

### Factors influencing IPC Compliance among healthcare workers

Out of 195 HCWs, 79 (41%) reported not being vaccinated against Hepatitis B infection, with 112(57.4%) and 107(54.9%) ever experiencing blood/body fluids exposure and needlestick injury, respectively. Despite the majority, 63.9%(n=124/195) of HCWs confirmed the presence of a full working IPC committee, 51(26.2%) have not received IPC supportive supervision at any period of the year while 59.5%, 45.1%, and 33.9% had not received any IPC training, received no motivation on IPC performance, and did not attend any IPC meeting in last year respectively. Table 4 shows the factors influencing IPC compliance among HCWs.

**Table 4:** Factors influencing IPC compliance among healthcare workers (N=195)

| Variable | Frequency | Percent* |
| --- | --- | --- |
| **Needlestick Injury** |  |  |
| Yes | 107 | 54.9 |
| No | 88 | 45.1 |
| **Exposed to blood/body fluids** |  |  |
| Yes | 112 | 57.4 |
| No | 83 | 42.6 |
| **Hepatitis B Vaccination** |  |  |
| Yes | 116 | 59.5 |
| No | 79 | 40.5 |
| **IPC trainings** |  |  |
| No training | 116 | 59.5 |
| Trained | 79 | 40.5 |
| **Motivation in the Workplace** |  |  |
| No motivation | 88 | 45.1 |
| Motivated | 107 | 54.9 |
| **Frequency of attending IPC training** |  |  |
| Monthly | 17 | 8.7 |
| Quarterly | 46 | 23.6 |
| Annually | 66 | 33.8 |
| Never | 66 | 33.9 |
| **Frequency of IPC supportive supervision** |  |  |
| Monthly | 49 | 25.1 |
| Quarterly | 29 | 14.9 |
| Annually | 66 | 33.9 |
| No supervision | 51 | 26.2 |
| **Presence of a full-time working IPC committee** |  |  |
| Yes | 124 | 63.6 |
| No | 71 | 36.4 |

\*Column percentages

### Factors associated with compliance with IPC practices among healthcare workers

Table 5 presents the results of a modified Poisson regression analysis examining factors associated with compliance with IPC practices among HCWs at KRRH. In the bivariate analysis, eight variables met the criteria for inclusion in the multivariable model: profession, needle-stick injury, exposure to body fluids, hepatitis B vaccination, IPC training, workplace motivation, frequency of IPC supportive supervision, and the presence of an IPC committee. After adjusting for potential confounders in the multivariable analysis, five variables remained significantly associated with IPC compliance: profession, exposure to body fluids, workplace motivation, frequency of IPC supportive supervision, and the presence of an IPC committee

**Table 5:** Factors associated with IPC compliance among HCWs (N=195)

| Variable | Compliance |  | P-value | CPR (95%CI) | P-value | APR (95% CI) | P<br>value |
| --- | --- | --- | --- | --- | --- | --- | --- |
|  | Non<br>compliance | Compliance |  |  |  |  |  |
| **Age (years)** |  |  |  |  |  |  |  |
| 18-30 | 63(65) | 34(35) | 1.599 | 1 |  | - |  |
| 31-40 | 48(58.3) | 35(41.7) |  | 1.19(0.82-1.72) | 0.362 |  |  |
| 41-50 | 7(50.0) | 7(50.0) |  | 1.42(0.78-2.580) | 0.239 |  |  |
| **Professions** |  |  |  |  |  |  |  |
| Nurse | 72(53.3) | 63(46.7) | **0.002** | 1 |  | 1 |  |
| Doctor/Clinician | 36(83.7) | 7(16.3) |  | 0.39(0.20-0.74) | **0.004** | 0.32(0.19 -0.57) | **<0.001** |
| Laboratory Technician | 11(64.7) | 6(35.3) |  | 0.73(0.38-1.43) | 0.363 | 0.68(0.34 -1.39) | 0.300 |
| **Sex** |  |  |  |  |  |  |  |
| Female | 51(62.2) | 31(37.8) | 0.081 | 1 |  | - |  |
| Male | 68(60.2) | 45(39.8) |  | 1.06(0.74-1.51) | 0.777 |  |  |
| **Needlestick Injury** |  |  |  |  |  |  |  |
| No | **61(69.3)** | **27(30.7)** |  | **1** |  | **1** |  |
| Yes | 55(51.4) | 52(48.6) | **0.011** | 1.58(1.09- 2.29) | **0.015** | 1.26(0.90- 1.76) | 0.176 |
| **Exposed to blood/body fluids** |  |  |  |  |  |  |  |
| No | 60(72.3) | 23(27.7) |  | 1 |  | 1 |  |
| Yes | 56(50.0) | 56(50.0) | **0.002** | 1.80(1.22- 2.68) | **0.003** | 1.55(1.095 -2.19) | **0.013** |
| **Hepatitis B Vaccination** |  |  |  |  |  |  |  |
| No | 36(45.6) | 43(54.4) |  | 1 |  | 1 |  |
| Yes | 80(69.0) | 44(31.0) | **0.001** | 0.57(0.41- 0.80) | **0.001** | 0.81(0.59- 1.11) | 0.185 |
| **IPC trainings** |  |  |  |  |  |  |  |
| No training | 77(66.4) | 39(33.6) | **0.018** | 1 | **0.017** | 1 |  |
| Received Training | 39(49.4) | 40(50.6) |  | 1.50(1.08- 2.11) |  | 1.27(0.95- 1.72) | 0.112 |
| **Motivation at the Workplace** |  |  |  |  |  |  |  |
| No | 64(72.3) | 24(27.3) | **0.001** | 1 |  | 1 |  |
| Yes | 52(48.6) | 55(51.4) |  | 1.89(1.28 -2.78) | **0.001** | 1.43(1.02- 2.02) | **0.040** |
| **Frequency of IPC supportive supervision** |  |  |  |  |  |  |  |
| No supervision | 41(80.4) | 10(19.6) |  | 1 |  | 1 |  |
| Monthly | 25(51.0) | 24(49.0) |  | 2.50(1.34 -4.67) | **0.005** | 1.40(0.76 -2.57) | 0.284 |
| Quarterly | 21(72.4) | 8(27.6)) | **<0.001** | 1.41(0.62- 3.17) | 0.410 | 0.89(0.42 -1.86) | 0.754 |
| Annually | 29(43.9) | 37(56.1) |  | 2.85(1.57 -5.19) | **0.001** | 1.92(1.09 -3.38) | **0.023** |
| **Presence of the IPC committee** |  |  |  |  |  |  |  |
| No | 54(76.1) | 17(23.9) |  | 1 |  | 1 |  |
| Yes | 62(50.0) | 62(50.0) | **<0.001** | 2.09(1.33-3.28) | **0.001** | 1.61(1.07- 2.40) | **0.021** |

**Adjusted for:** Profession, needle stick injury, exposure to blood/body fluids, Hepatitis B vaccination, IPC training motivation at the workplace, frequency of IPC supportive supervision, and presence of IPC committee.

Doctors and clinicians were 0.32 times less likely to comply with IPC practices compared to nurses (APR = 0.32, 95% CI: 0.19 – 0.57, p <0.001). This difference was statistically significant. HCWs who had ever encountered body fluid exposure were 1.55 times more likely to comply with IPC practices compared to those who had not been exposed, and this was statistically significant (APR = 1.55, 95% CI = 1.10 - 2.19, p = <0.013). HCWs who receive IPC motivation at the workplace were 1.43 times more likely to comply with IPC practices compared to those who don’t receive IPC motivation, and this difference was statistically significant (APR =1.43, 95% CI = 1.02-2.02, p = 0.040).

HCWs who received IPC supportive supervision annually exhibited a higher likelihood of adhering to IPC practices, in comparison to HCWs with no supportive supervision. This increased adherence was significantly associated with optimal compliance, as indicated by the adjusted prevalence ratio (APR =1.92, 95% CI =1.09 - 3.38, p = 0.023). Likewise, HCWs who confirmed the presence of the IPC committee were 1.61 times more likely to comply with IPC practices compared to those who reported the absence of the IPC committee, and this was statistically significant (APR = 1.61, 95% CI = 1.07-2.40, p = 0.021). There was no significant association between other factors, such as needle stick injury, hepatitis B vaccination, and IPC training.

### Availability of IPC supplies at KRRH departments

The availability of IPC supplies plays a crucial role in how HCWs implement IPC practices in a health facility. During the observation of IPC supplies at KRRH, nine departments (Emergency, neonatal, Intensive care unit, paediatric, medical, surgical, theatre, laboratory, and maternity) were assessed for the availability of IPC supplies using an observational checklist. These departments were selected because they represent areas where IPC practices are most critical for preventing HAI.

Based on the observation (Table 6), the most available IPC supplies in the department were hand hygiene items (100%), injection safety (100%), sterilization (100%), and waste management (100%). The majority of departments (7) had PPE supplies like face masks, disposable gloves, and gowns/aprons/lab coats (100%), while two departments were missing protective eyewear. Nearly all departments had functional and clean latrines, while five departments had no soap, and four departments lacked constantly running water for hand washing in the latrine.

**Table 6:**
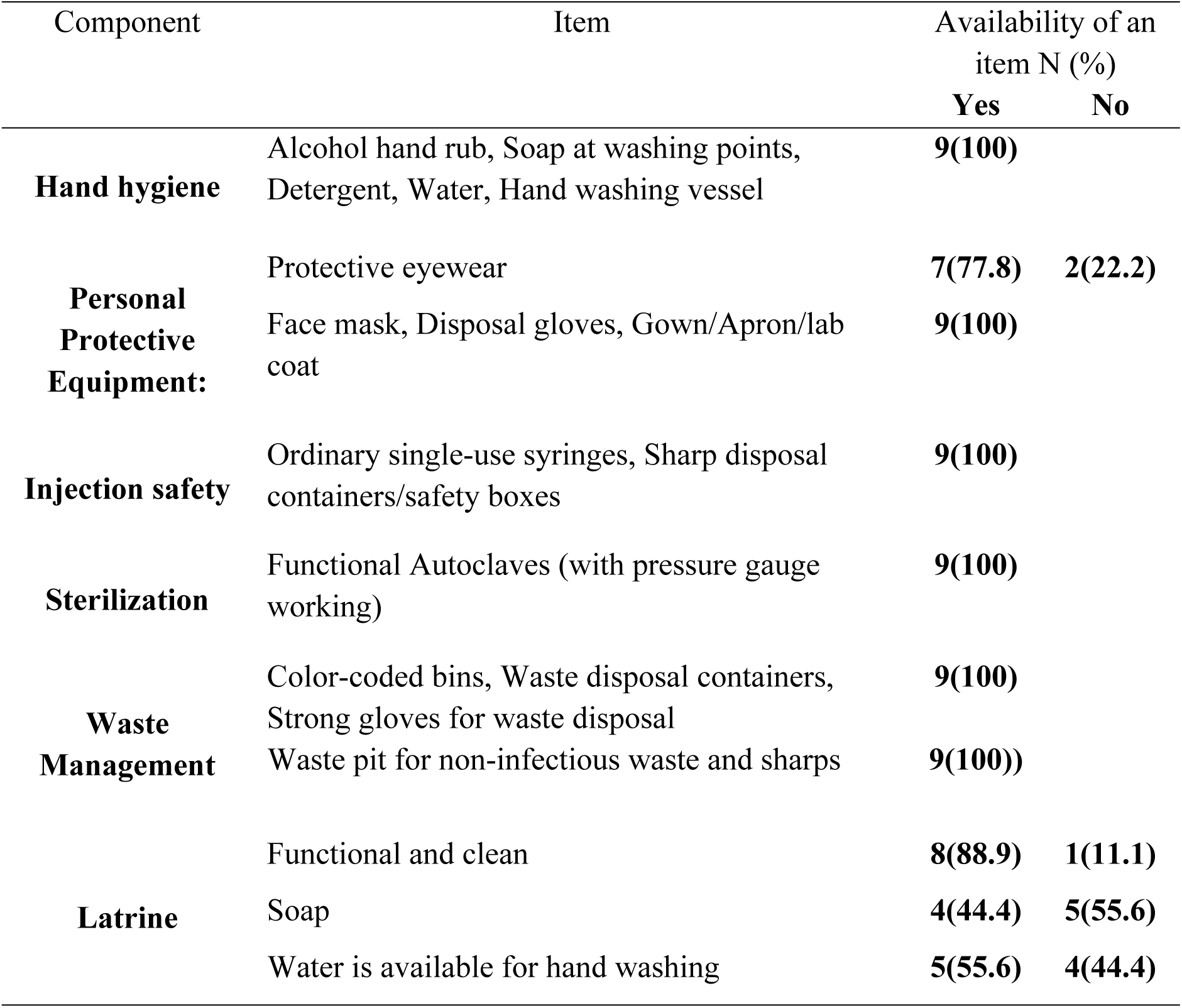
Availability of IPC supplies at KRRH departments via an observational checklist.

## Discussion

This study aimed to assess overall compliance with Infection Prevention and Control Practices (IPC) and their associated factors among HCWs at KRRH.

The low overall compliance results observed in this study align with related studies conducted in River State (59%), Rwanda (64.5%), Lethoto (63.6%), Ethiopia (65%), and parts of Tanzania, such as Songwe region (66%) and St. Francis Regional Referral Hospital in Tanzania (54%) [28,31–33]. This may be attributed to ongoing challenges, such as inadequate PPE supplies, a lack of motivation, and negative attitudes toward IPC practices, which are experienced in many health facilities in lower- and middle-income countries, including Tanzania. However, our study results are higher than those in similar studies conducted in Iran (34%), Bangladesh (36%), Ghana (45.1%), and Nigeria (50.8%) [32]. This difference could stem from variations in study periods, sample sizes, design, and social disparities between countries. The overall low compliance with IPC practices highlights the urgent need for policymakers, the management team, and healthcare workers at KRRH to strengthen strategies that promote the implementation of IPC practices and increase overall compliance.

More than half of HCWs in the current study do not use PPE, such as wearing a surgical mask or a combination of goggles, a face shield, and an apron whenever there is a risk of splash exposure. In addition, the majority of HCWs rely on water alone for hand washing despite the availability of some IPC resources such as alcohol hand rub, soap at washing points, and detergent. Similar results have been reported in Ethiopia, Uganda, and India, where poor PPE use and inadequate hand hygiene were linked to inconsistent PPE supply, discomfort, and perceived low risk [34–36]. The observed finding in this study may be due to the existence of an inconsistent supply of PPEs and inadequate enforcement of IPC protocols, as explained by the majority of HCWs in qualitative results.

In contrast, the Kenyan study by Brown et al in health facilities found that HCWs followed IPC practices by wearing full PPE, mainly due to ongoing IPC training and strict supervisory checks [37]. This could be due to ongoing strict supervision, as the study took place during the COVID-19 pandemic era, with the ongoing Ebola outbreak across the border in Uganda. These findings suggest that even in a resource-constrained environment, sustained training and continuous supervision can significantly improve compliance with IPC measures.

In this study, doctors and clinicians showed notably lower compliance with IPC compared to nurses. Similar results have been reported by other studies in the United Kingdom, Bangladesh, Australia, and Songwe, Tanzania [9,33,38,39]. Plausible explanations for the current study may be associated with multiple factors. For nurses, for example, the inclusion of additional IPC courses in their training programmes may contribute by equipping them with basic knowledge before being enrolled in service. Also, a shortage of doctors’ results in an excessive workload and a corresponding decline in participation in in-service IPC courses. These observations are in line with qualitative findings in the current study, which revealed that a high volume of patients with understaffing increased the likelihood of HCWs not complying with IPC practices. This highlights the urgent need for targeted IPC educational training among healthcare workers, irrespective of cadres, as well as workload management strategies, to bridge the compliance gap and improve overall infection control outcomes at KRRH and similar healthcare settings.

Healthcare workers who were exposed to body fluids were 1.5 times more likely to comply with IPC practices than those who were not exposed, implying that heightened risk perception is a strong driver of compliance. These findings are in line with other studies conducted in Ethiopia, which showed that HCWs with higher perceived risk had markedly better compliance with IPC guidelines [40]. This significant observation in this study underscores the importance of strengthening risk perception through continuous education and awareness, and acts as a call to action for KRRH management and HCWs to reinforce compliance with IPC practices even in the absence of immediate exposure risks. In contrast to this observation is the study conducted in Ghana, which showed that HCWs who encountered blood exposure had lower compliance with the IPC guidelines [41]. The difference in findings may be due to differences in institutional policies and the coexistence of a weak safety culture in most resource-limited facilities. However, exposure alone is insufficient without system support [42].

Furthermore, the current study demonstrated that, despite having no significant association, IPC training is important in enhancing knowledge of IPC protocols and practices, as revealed by HCWs who attended IPC training, showing a 1.2 times likelihood of IPC compliance compared to those who had not attended training. These findings are consistent with related studies in Ethiopia, Ghana, and parts of Tanzania [32,39,43,44] which highlights the need for the Ministry of Health, KRRH management, and HCWs to strengthen regular IPC training to sustain and improve compliance. The observed training culture in this study, where knowledge is cascaded from those trained by the Ministry of Health to their colleagues, reflects a peer-learning model that has been reported to increase reach and sustainability of IPC practices in resource-limited settings.

Healthcare workers who received IPC motivation at the workplace in the current study were more likely to comply with IPC practices than those not motivated. These findings are in line with a study conducted by Houghton et al, who reported that self-motivation among HCWs by perceiving IPC value of protecting themselves, their families, or their patients is a behavioural change motive towards IPC compliance [45]. Moreover, a supportive workplace culture and managerial backing further reinforce compliance among motivated staff [46]. Thus, intervention involving self-motivation, including incentives, non-monetary, and recognition, can significantly influence IPC compliance among HCWs at KRRH and similar settings.

Furthermore, monitoring of the IPC practices via supportive supervision plays a vital role in enhancing compliance with the IPC practices. This observation forms the basis for the current study. Findings revealed HCWs who received IPC supportive supervision were more likely to comply with IPC practices compared to those with no supportive supervision. These findings align with findings from studies done in Gambia [45,47]. During supportive supervision, IPC activities are monitored, implementation gaps are identified, and addressed. Therefore, a continuous and thorough monitoring system for IPC practices among HCWs should be established. It should be carried out regularly and in a non-punitive way, focusing on identifying areas for improvement and providing constructive feedback to HCWs. This approach will not only help identify compliance gaps but also motivate HCWs to consistently follow IPC practices.

Additionally, the current study revealed that HCWs who confirmed the presence of the IPC committee were more likely to comply with IPC practices compared to those who reported the absence of the IPC committee. A similar study done across health facilities in the Kampala region during the COVID-19 era emphasized that for the HCWs to comply with IPC practices, there is a need for IPC committee activities and work plans to be supported by the budget lines [48]. This reflects that the IPC committee influences HCWs to adhere to IPC practices via its activities, such as routine monitoring, providing feedback on IPC strategies, and the provision of reminders on IPC.

### Strengths and Limitations of the Study

The strength of this study lies in its approach of using multiple data collection tools: questionnaires and observational checklists to gather data directly from the required participants, giving valuable insights for healthcare workers, health administrators, and policymakers. Additionally, the study identified several key findings that need further research with a larger sample and a more robust design, such as a longitudinal study, direct observation, and qualitative perspectives.

Despite the strength, this study has certain limitations. Initially, being a cross-sectional study makes it difficult to infer causality. Secondly, the measurement of IPC compliance relied on self-reported responses from the participants, which may introduce the possibility of social desirability bias influencing the accuracy of the data. Furthermore, only participants directly involved in patient care were included in the study, thus limiting generalizability. Although these limitations may have introduced some bias or reduced the generalizability of the findings, their overall impact on the study’s conclusions is considered minimal.

## Conclusion and recommendation

Overall compliance with IPC practices among HCWs was suboptimal, with only 39% achieving the recommended level of >80%. Compliance was positively influenced by exposure to body fluids, workplace motivation, supportive supervision, and the presence of an IPC committee. While essential supplies such as hand hygiene items, PPEs, and waste management items were available, gaps remained, particularly the lack of water and soap in latrines.

To reduce the HAI burden, healthcare workers, regardless of their profession, should integrate the IPC practices as the core component in healthcare delivery by utilizing the available IPC supplies and staying updated with IPC knowledge. Healthcare facilities should strengthen the IPC activities within the IPC committees and address the IPC resource gaps as necessary interventions for improvement of HCWs’ compliance with IPC practices. Additionally, health management teams at the national, regional, and facility levels should ensure consistent preparation and scale-up refresher IPC trainings towards all HCWs and encourage data-driven IPC decision-making by investing in research and surveillance in order to leverage quality improvement activities in healthcare facilities and improve universal compliance.

## Data Availability

All relevant data are within the manuscript and its Supporting Information files

## Acknowledgements

We express our heartfelt thanks to all individuals who participated in the study: respondents, data collectors, and administrative officials

## List of observations

AMR: Antimicrobial Resistance
HAIs: Hospital-Acquired Infections
HBV: Hepatitis B Virus
HH: Hand Hygiene
IPC: Infection Prevention and Control
KRRH: Katavi Referral Regional Hospital
PPEs: Personal Protective Equipment

## Authors’ Contributions

**Conceptualization:** Cesilia Charles, Lutengano Mkonongo, David Masanja, Pendo Edward, Frank Elisha, Deogratius Banuba

**Data curation:** Emmanuel Amsi, Elly Ambilikile, Avent Kalumiana

**Formal analysis:** Joshua Mollel, Baraka Nkondo, Philipo Mwita

**Investigation:** Edward Bucheye, Abel Nyika

**Methodology:** Cesilia Charles, Lutengano Moshi, Damiani Maruba

**Project administration:** Frank Elisha, Deogratius Banuba

**Resources:** Deogratius Banuba

**Software:** Cesilia Charles, Lutengano Mkonongo, Joshua Mollel

**Supervision:** Frank Elisha, Deogratius Banuba

**Validation:** Lutengano Mkonongo

**Writing original draft:** Cesilia Charles, Lutengano Mkonongo

**Writing review and editing:** Cesilia Charles, Radenta Bahegwa, Bernard Njau, Nathanael Sirili

